# Macula structural and vascular differences in glaucoma eyes with and without high axial myopia

**DOI:** 10.1101/2021.09.02.21263045

**Authors:** Jasmin Rezapour, Christopher Bowd, Jade Dohleman, Akram Belghith, James A. Proudfoot, Mark Christopher, Leslie Hyman, Jost B. Jonas, Rafaella C. Penteado, Sasan Moghimi, Huiyuan Hou, Massimo A. Fazio, Robert N. Weinreb, Linda M. Zangwill

**Author notes:** Corresponding author: Linda M. Zangwill, 9500 Gilman Drive, La Jolla, CA 92093-0946, Shiley Eye Institute/Hamilton Glaucoma Center, Viterbi Family Department of Ophthalmology, University of California, San Diego. **Precis** In glaucoma eyes, macula ganglion cell thickness measures were significantly associated with severity of glaucoma but not axial length suggesting that macula OCT parameters may be useful in detecting glaucoma in eyes with high myopia.

## Abstract

**Aims:** To assess the thickness of various retinal layers, and the superficial vessel density (sVD) in the macula of glaucomatous eyes and their associations with axial length (AL) and visual field mean deviation (VFMD) to identify parameters useful for glaucoma management in myopic eyes.

**Methods:** 248 glaucoma patients (401 eyes) participating in the Diagnostic Innovations in Glaucoma Study observational cohort representing 3 axial myopia groups (non-myopia: n=146 eyes; mild myopia: n=208 eyes; high myopia (AL>26 mm): n=47 eyes) who completed macular OCT and OCT-Angiography imaging were included. The cross-sectional associations of AL and VFMD with the thickness of the ganglion cell inner plexiform layer (GCIPL), macular retinal nerve fiber layer (mRNFL), ganglion cell complex (GCC), sVD and macular choroidal thickness (mCT) were evaluated.

**Results:** Thinner Global GCIPL and GCC were significantly associated with worse VFMD (R^2^=35.1%; and R^2^=33.4%; respectively p<0.001), but not with AL (all p>0.350). Thicker mRNFL showed a weak association with increasing AL (R^2^=3.4%; p=0.001) and a positive association with VFMD (global R^2^=20.5%; p<0.001). Lower sVD was weakly associated with increasing AL (R^2^=2.3%; p=0.016) and more strongly associated with more severe glaucoma VFMD (R^2^=31.8%; p<0.001). Thinner mCT was associated with increasing AL (R^2^=17.3% p<0.001) and not associated with VFMD (P=0.262). mRNFL was thickest while mCT was thinnest in all sectors of high myopic eyes.

**Conclusions:** GCIPL and GCC thinned with increasing severity of glaucoma but were not significantly associated with axial length. GCIPL and GCC thickness may be useful clinical parameters to identify glaucoma in myopic eyes.

## Introduction

With its potential vision threatening risk and with its prevalence increasing globally, myopia, especially high myopia, has become a major concern around the world.^1^ Although optical coherence tomography (OCT) based measurements of peripapillary retinal nerve fiber layer (pRNFL) thickness can accurately discriminate between healthy and glaucomatous eyes,^2^ there is concern that in myopic eyes (and especially in high myopic eyes) the diagnostic accuracy of OCT measures is decreased. Optic disc changes in myopic eyes such as morphologic changes in the parapapillary region and optic disc enlargement pose significant challenges to the use of optical imaging and clinical optic disc evaluation to detect and monitor glaucoma (Figure 1).^1,3^ This is due in part to difference in the regional arrangement of the peripapillary retinal nerve fibers between myopic eyes and emmetropic eyes that may result in sectoral values incorrectly classified as outside normal limits by instrument-specific software analysis in healthy myopic eyes.^4 5^

**Figure 1:**
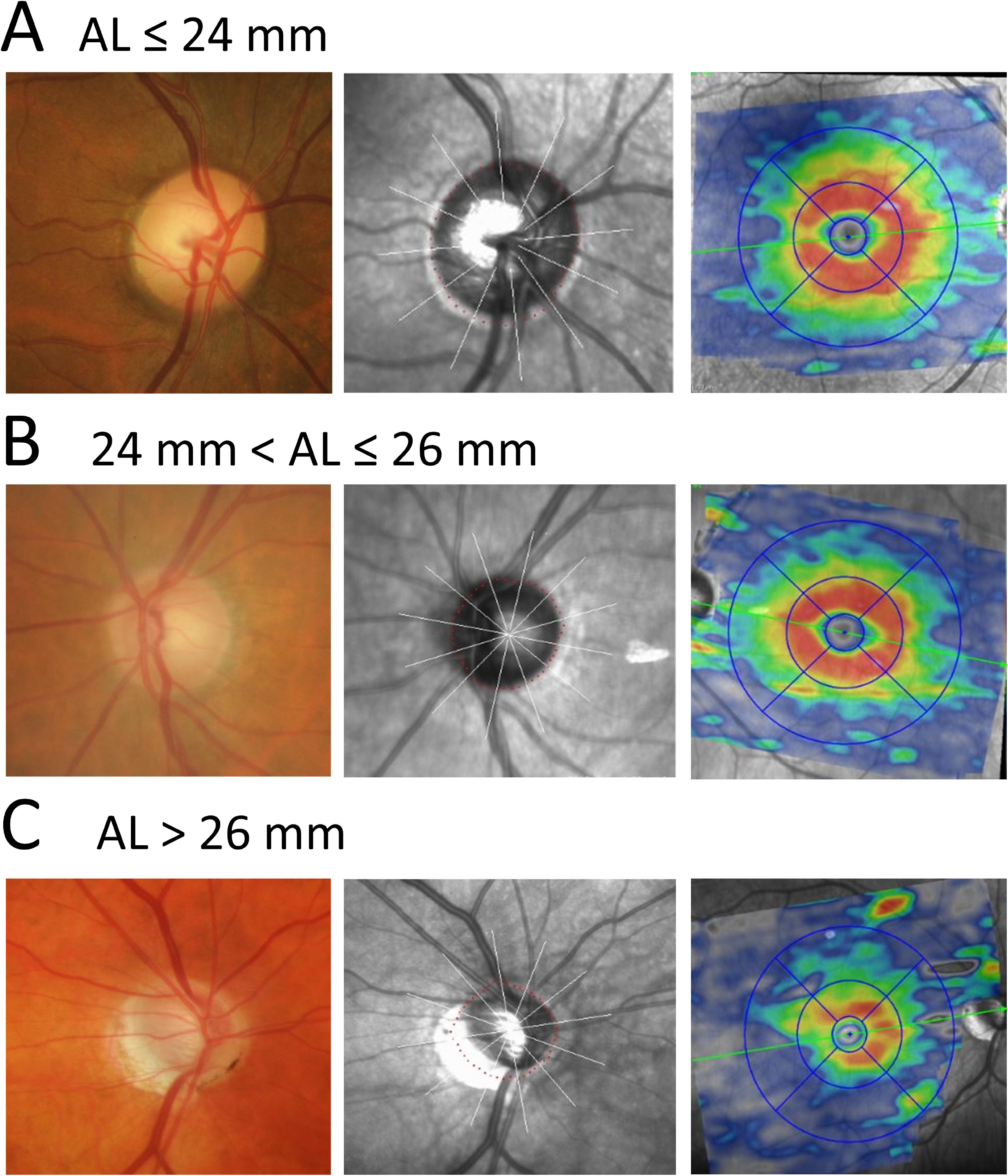
Optic disc photograph (left), optical coherence tomography optic nerve head en face image (middle) and optical coherence tomography macula posterior pole image (right) of an eye with (A) no axial myopia (axial length = 23.8 mm), (B) mild axial myopia (axial length = 24.7 mm and (C) high axial myopia (axial length = 29 mm).

Approximately 50% of the retinal ganglion cells are concentrated within 10 degree of the fovea^6^ making the macula an useful region for diagnosing optic neuropathies including glaucoma, especially in myopic eyes because myopic axial elongation primarily affects the optic nerve head region. Previous studies have reported that early glaucomatous damage can be detected in the macula region^7^ and that measurements of the ganglion cell inner plexiform layer (GCIPL) can be used for detecting glaucoma in highly myopic eyes.^8-12^ However, little information is available about differences in the topographic distribution of the thickness of the various macular retinal layers and the retinal vessel density in glaucomatous eyes with and without myopia. Sectoral measurements of the underlying macular vasculature may offer additional insight into differences in glaucomatous eyes with and without myopia.

Several studies using OCT-Angiography (OCTA) have demonstrated a strong relationship between macular capillary density and the severity of glaucoma.^13 14^ Furthermore recent studies have reported the peripapillary choroid to be thinner in highly myopic eyes compared to non-myopic eyes.^15 16^ However, few studies have assessed macular choroidal thickness in highly myopic eyes^17 18^ and to date, to the best of our knowledge, no study has documented the local distribution of macular choroidal thickness in glaucomatous eyes with and without high myopia.

The purpose of this study was to characterize the local distribution of GCIPL, GCC, macular retinal nerve fiber layer (mRNFL), choroidal thickness and vessel density in glaucoma eyes with and without axial myopia. By better understanding how the topographic distribution of these parameters varies with axial length and severity of disease, macula parameters that may be useful for detecting and monitoring glaucoma in myopic eyes can be identified.

## Methods

### Study Sample

This cross-sectional study included all glaucoma patients enrolled in the University of California, San Diego Diagnostic Innovations in Glaucoma Study (DIGS; clinicaltrials.gov identifier NCT00221897) with available axial length measurements and good quality macula OCT scans acquired between 2015 and 2020. The study was approved by the institutional review board of the University of California San Diego and according to the tenets of the Declaration of Helsinki written informed consent was obtained from all patients. As described previously;^19^ participants underwent a complete ophthalmologic examination including assessment of refractive error, axial length measurement (IOLMaster, Carl Zeiss Meditec, Dublin, CA), visual field testing, simultaneous stereophotography of the optic disc and macula, and macular OCT and OCTA imaging. Study participants were ≥18 years with best-corrected visual acuity ≥20/40 and open anterior chamber angles at baseline.

Visual field (VF) testing was performed using the standard Humphrey Field Analyzer 24-2 Swedish interactive thresholding algorithm. Repeatable glaucomatous VF damage was defined as the presence of glaucomatous optic nerve head (ONH) damage based on masked assessment by two trained observers and glaucomatous VF damage.^19^ ONH stereophotographs of highly myopic eyes were graded for glaucoma by two experts (CB and JR) after training with a senior consultant (JBJ). Diagnosis was defined by consensus between the two graders and adjudication by the senior consultant in case of disagreement.

### Myopia definition

Because a change in refractive error can occur after refractive or cataract surgery, myopia was classified by axial length into the following 3 groups.

- No myopia: axial length ≤ 24.0 mm
- Mild myopia: 24.0mm < axial length ≤ 26.0 mm
- High myopia: axial length > 26.0 mm

### Optical coherence tomography and optical coherence tomography angiography imaging

OCT imaging of the macula was performed with the Spectralis OCT (version 6.10; Heidelberg Engineering Inc, Heidelberg, Germany). Details of this instrument have been previously described.^14^ Macula horizontal posterior pole (p-Pole) scans covering an area of 30° × 25° (6 × 6 mm) were obtained. GCIPL, mRNFL and GCC (GCIPL + mRNFL) thickness measurements were generated from each retinal layer from the central 1-, 3-, and 6-mm circles as inner rings (1- and 3-mm circle) and outer rings (3- and 6-mm circle) according to the Early Treatment Diabetic Retinopathy Study defined sectors (temporal, superior, nasal, and inferior).

OCTA imaging of the macula was performed with the Avanti AngioVue OCT system (version 2017.1.0.151; Optovue, Inc., Fremont CA, USA).^20^ Macular whole image vessel density was calculated on a 3 × 3 mm^2^ field macula scan (304 B-scans × 304 A-scans per B-scan) centered on the fovea. Whole image vessel density of the temporal, superior, nasal, and inferior sectors were reported. Macular parafoveal superficial VD (sVD) was calculated within an annulus centered on the fovea, with an inner diameter of 1 mm and an outer diameter of 2.5 mm.

All images were reviewed by the Imaging Data Evaluation and Analysis (IDEA) Reading Center for image quality, and accurate segmentation of the mRNFL, ganglion cell layer (GCL), and inner plexiform layer (IPL). The automated Spectralis software segmentation was manually corrected if needed, according to the standard IDEA Reading Center protocols.^19^

### Choroidal thickness measurement using deep learning

As choroidal thickness is not available from standard software, custom deep learning-based software was developed to automatically measure mCT.^21^ A trained grader (JR) manually segmented the Bruch’s Membrane (BM) / RPE complex and the posterior boundary of the choroid in 120 p-Pole scans in the SPX software (version 1.9.204.0; Heidelberg Engineering Inc, Heidelberg, Germany) in a subset of 20 eyes, which was used as ground truth to train a deep convolutional neural network model (BCDU-Net).^22^ Two thousand two hundred fifty seven scans (753 eyes) with automated choroid segmentation were reviewed for accuracy (JR). The overall performance of the deep learning algorithm for segmenting the choroid was very good with 400/401 (99.8%) eyes included (no myopia: 146/146 (100%), mild myopia: 208/208 (100%) and high myopia: 46/47 (97.9%)).

Macular choroidal thickness (mCT) was obtained for the inner and outer rings of the macular p-Pole scans defined above. Each ring was subdivided into temporal, superior, nasal, and inferior sectors and global and sectoral choroidal thickness was calculated. The MCT was defined as the perpendicular distance between the posterior border of BM / retinal pigment epithelium (RPE) complex and the posterior boundary of the choroid.

### Statistical Analyses

Data is presented as mean (95% confidence interval (CI)) and count (percentage) for continuous and categorical variables, respectively. Patient and eye characteristics were compared across myopia groups using analysis of variance (ANOVA) and chi-squared tests for continuous and categorical patient-level variables (respectively) and linear mixed effects models for continuous eye-level variables, with a random intercept to account for within-patient correlation. Univariable and age and VFMD adjusted multivariable models were applied to evaluate the association between axial length and ocular parameters. P-values less than 0.05 were considered statistically significant. All statistical analyses were performed using R (version 3.6.3).

## Results

Four-hundred-one glaucoma eyes of 248 patients were included with 146 eyes (87 patients) in the non-myopic group, 208 eyes (125 patients) in the mild myopic group, and 47 highly myopic eyes (36 patients) (Table 1). All p-values are reported as age-adjusted.

**Table 1:**
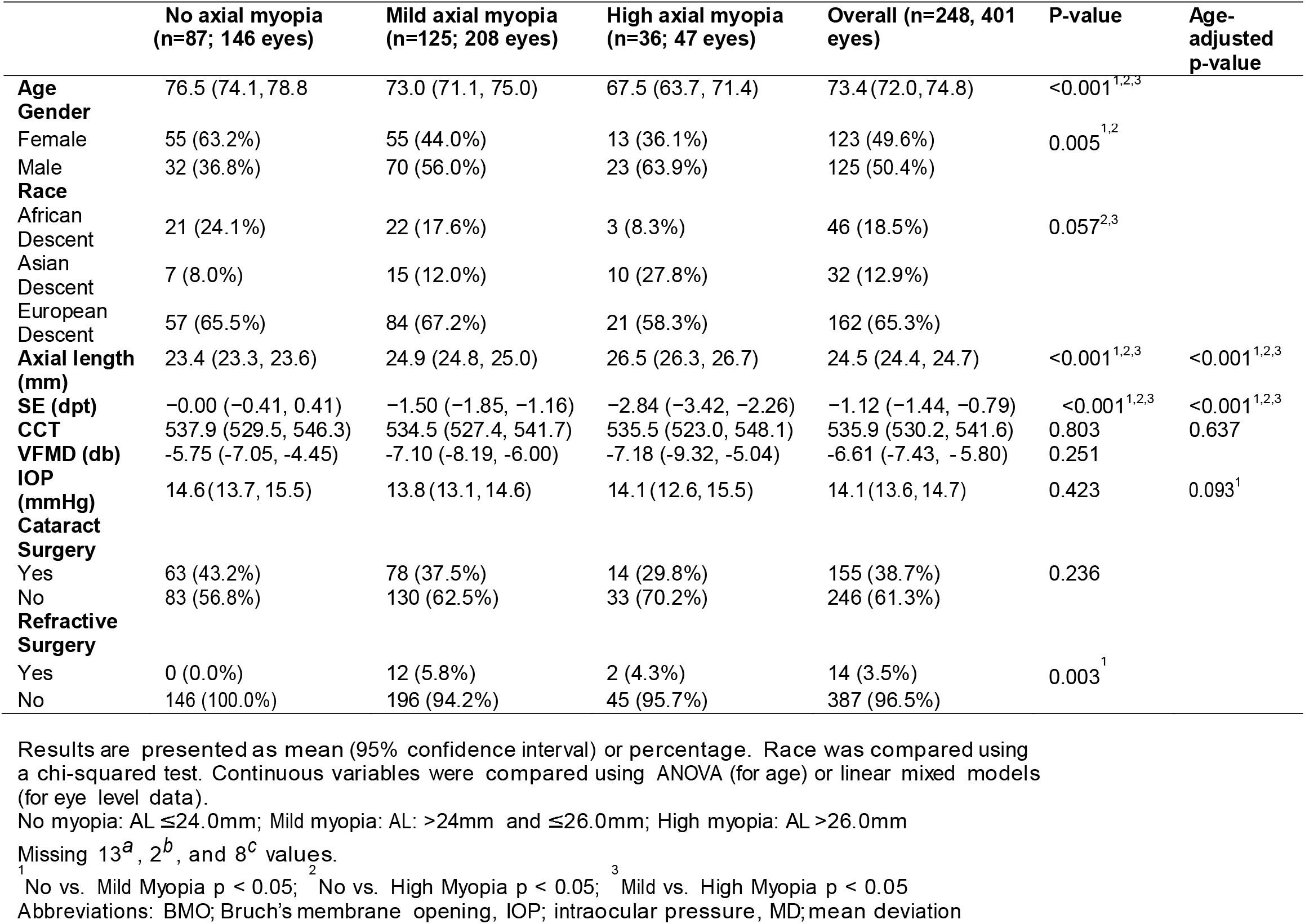
Glaucoma patient and eye characteristics by myopia group.

The participants in the high myopia group were significantly younger (mean [95% CI]) 67.5 [63.7, 71.4] years) than the members of the mild (73.0 [71.1, 75.0] years) myopic group and the non-myopic individuals (76.5 [74.1, 78.8] years) groups (p<0.001). There was a trend of a higher proportion of individuals of Asian descent in the high myopia group compared to the no-myopic and the mild myopic group (p=0.06).

There was no significant difference in intraocular pressure (p=0.09), central corneal thickness (p=0.64) and BMO area (p=0.51) among the three groups (Table 1). Non-myopes tended (p=0.131) to have less severe glaucoma than the mild myopic group and the high myopic group (mean visual field mean deviation (MD) -5.75 dB, -7.10 dB and -7.18 dB, respectively).

### Macular Thickness Measurements

A total of 43 eyes were excluded from the analysis for not meeting image quality criteria (Spectralis quality score >15 dB or segmentation failure) with 14/140 (10.1%), 12/220 (5.5%) and 17/64 (26.6%) eyes excluded from the no-, mild- and high-axial myopia groups, respectively. Macular thickness measures are presented in Figure 2 and Supplemental Table 1.

**Figure 2:**
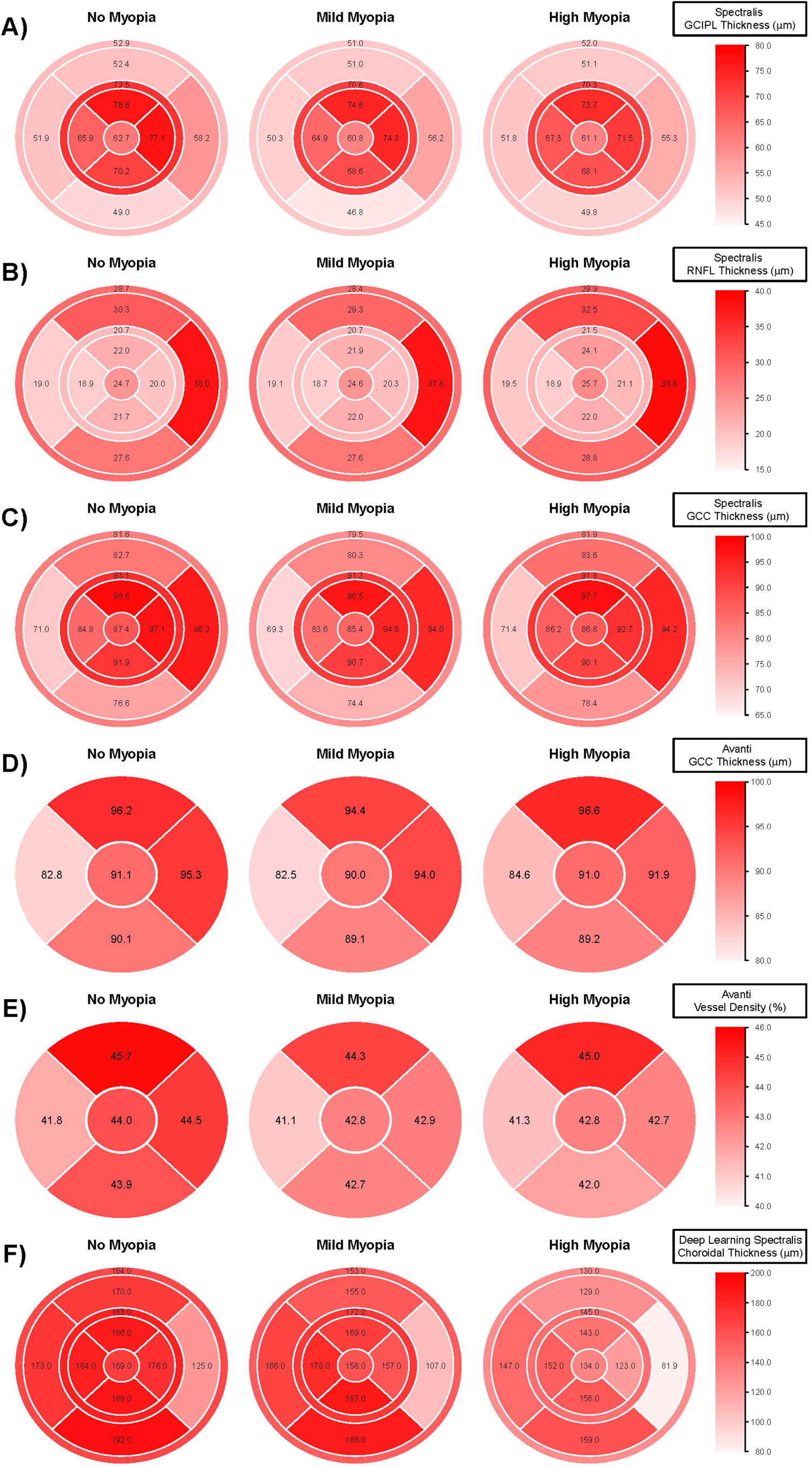
Sectoral and global thickness distribution of the Spectralis GCIPL thickness (1A), Spectralis macular RNFL thickness (1B), Spectralis GCC thickness (1C), Avanti GCC thickness (1D), Avanti macular vessel density (1E) and Spectralis macular choroidal thickness (1F) in non-myopic, mild myopic and highly myopic glaucoma eyes. Abbreviations: GCC; ganglion cell complex, GCIPL; ganglion cell inner plexiform layer, RNFL; retinal nerve fiber layer

### Associations with Axial Length

There were no statistically significant associations between global and sectoral GCC or GCPIL thickness measurements and axial length except for a weak association of the GCIPL outer nasal sector (R^2^=1.9%, p=0.016). All mRNFL measurements, except for the inner temporal and outer inferior sector were significantly (all p<0.024) but relatively weakly (all R^2^ < 5%) associated with axial length. We found weak associations between global and sectoral vessel density measurements and axial length (all R^2^≤3.2%, all p<0.05) and stronger associations between choroidal thickness measures and axial length (R^2^ range: 9.6%-19.3%, all p<0.001) (Table 2).

**Table 2.** Ocular associations with axial length

|  | Patients (eyes) | Univariable Regression |  | Multivariable Regression |  |
| --- | --- | --- | --- | --- | --- |
|  |  | Estimate | R<sup>2</sup> (p-value) | Estimate | R<sup>2</sup>(%) (p-value) |
| **Age** | N=248 (401) | -0.26 (-0.59, 0.07) | 0.1 (0.121) | -0.25 (-0.58, 0.07) | 0.1 (0.130) |
| **VFMD** | N=248 (401) | -0.35 (-0.97, 0.27) | 0.4 (0.272) | -0.53 (-1.17, 0.12) | 0.8 (0.110) |
| **Spectralis GCIPL (μm)** |  |  |  |  |  |
| Global | N=248 (401) | -0.29 (-1.21, 0.63) | 0.1 (0.531) | -0.41 (-1.19, 0.36) | 0.4 (0.299) |
| Inner ring | N=248 (401) | -0.32 (-1.58, 0.94) | 0.1 (0.618) | -0.36 (-1.42, 0.69) | 0.2 (0.498) |
| Outer ring | N=248 (401) | -0.26 (-0.92, 0.40) | 0.2 (0.443) | -0.45 (-1.05, 0.14) | 0.7 (0.136) |
| Inner temporal | N=248 (401) | 0.14 (-1.26, 1.54) | 0.0 (0.843) | 0.29 (-0.86, 1.43) | 0.1 (0.625) |
| Inner superior | N=248 (401) | -0.60 (-1.90, 0.71) | 0.3 (0.370) | -0.82 (-2.01, 0.37) | 0.6 (0.180) |
| Inner nasal | N=248 (401) | -0.61 (-1.96, 0.74) | 0.3 (0.380) | -0.76 (-1.99, 0.47) | 0.5 (0.226) |
| Inner inferior | N=248 (401) | -0.21 (-1.69, 1.26) | 0.0 (0.780) | -0.11 (-1.37, 1.15) | 0.0 (0.859) |
| Outer temporal | N=248 (401) | -0.20 (-1.01, 0.62) | 0.1 (0.637) | -0.20 (-0.91, 0.51) | 0.1 (0.577) |
| Outer superior | N=248 (401) | -0.21 (-0.92, 0.50) | 0.1 (0.567) | -0.47 (-1.14, 0.20) | 0.6 (0.170) |
| Outer nasal | N=248 (401) | -0.49 (-1.28, 0.30) | 0.5 (0.225) | -0.92 (-1.66, -0.18) | 1.9 (0.016) |
| Outer inferior | N=248 (401) | -0.18 (-0.84, 0.48) | 0.1 (0.595) | -0.27 (-0.90, 0.37) | 0.2 (0.411) |
| **Spectralis RNFL (μm)** |  |  |  |  |  |
| Global | N=248 (401) | 0.33 (-0.05, 0.71) | 0.9 (0.092) | 0.59 (0.24, 0.95) | 3.4 (0.001) |
| Inner ring | N=248 (401) | 0.32 (0.06, 0.59) | 1.8 (0.018) | 0.49 (0.22, 0.76) | 3.9 (<0.001) |
| Outer ring | N=248 (401) | 0.33 (-0.21, 0.87) | 0.5 (0.233) | 0.70 (0.21, 1.19) | 2.6 (0.005) |
| Inner temporal | N=248 (401) | 0.04 (-0.16, 0.24) | 0.1 (0.671) | 0.19 (-0.01, 0.39) | 1.0 (0.066) |
| Inner superior | N=248 (401) | 0.55 (0.18, 0.92) | 2.7 (0.004) | 0.74 (0.35, 1.12) | 4.4 (<0.001) |
| Inner nasal | N=248 (401) | 0.44 (0.09, 0.79) | 1.9 (0.014) | 0.62 (0.25, 0.98) | 3.4 (<0.001) |
| Inner inferior | N=248 (401) | 0.27 (-0.09, 0.63) | 0.7 (0.141) | 0.44 (0.09, 0.80) | 1.8 (0.015) |
| Outer temporal | N=248 (401) | 0.07 (-0.13, 0.28) | 0.2 (0.483) | 0.23 (0.03, 0.44) | 1.6 (0.024) |
| Outer superior | N=248 (401) | 0.44 (-0.28, 1.16) | 0.5 (0.231) | 0.83 (0.15, 1.51) | 1.8 (0.017) |
| Outer nasal | N=248 (401) | 0.60 (-0.23, 1.43) | 0.6 (0.160) | 1.14 (0.36, 1.93) | 2.7 (0.004) |
| Outer inferior | N=248 (401) | 0.17 (-0.57, 0.91) | 0.1 (0.654) | 0.59 (-0.09, 1.26) | 0.9 (0.088) |
| **Spectralis GCC (μm)** |  |  |  |  |  |
| Global | N=248 (401) | 0.03 (-1.19, 1.25) | 0.0 (0.964) | 0.18 (-0.88, 1.25) | 0.0 (0.737) |
| Inner ring | N=248 (401) | 0.00 (-1.41, 1.41) | 0.0 (0.998) | 0.12 (-1.10, 1.35) | 0.0 (0.844) |
| Outer ring | N=248 (401) | 0.07 (-1.04, 1.17) | 0.0 (0.909) | 0.25 (-0.75, 1.24) | 0.1 (0.625) |
| Inner temporal | N=248 (401) | 0.18 (-1.24, 1.61) | 0.0 (0.800) | 0.47 (-0.72, 1.66) | 0.2 (0.438) |
| Inner superior | N=248 (401) | -0.06 (-1.59, 1.48) | 0.0 (0.943) | -0.08 (-1.53, 1.36) | 0.0 (0.910) |
| Inner nasal | N=248 (401) | -0.16 (-1.68, 1.37) | 0.0 (0.840) | -0.14 (-1.58, 1.31) | 0.0 (0.852) |
| Inner inferior | N=248 (401) | 0.06 (-1.68, 1.80) | 0.0 (0.947) | 0.33 (-1.20, 1.85) | 0.1 (0.676) |
| Outer temporal | N=248 (401) | -0.12 (-1.05, 0.82) | 0.0 (0.805) | 0.03 (-0.80, 0.87) | 0.0 (0.940) |
| Outer superior | N=248 (401) | 0.23 (-1.07, 1.53) | 0.0 (0.725) | 0.36 (-0.87, 1.58) | 0.1 (0.569) |
| Outer nasal | N=248 (401) | 0.12 (-1.31, 1.55) | 0.0 (0.873) | 0.22 (-1.13, 1.56) | 0.0 (0.750) |
| Outer inferior | N=248 (401) | -0.02 (-1.30, 1.27) | 0.0 (0.978) | 0.31 (-0.88, 1.51) | 0.1 (0.608) |
| **Avanti GCC (μm)** |  |  |  |  |  |
| Whole image | N=204 (317) | -0.04 (-1.44, 1.36) | 0.0 (0.954) | 0.02 (-1.23, 1.26) | 0.0 (0.979) |
| Parafovea | N=204 (317) | -0.15 (-1.64, 1.33) | 0.0 (0.839) | -0.13 (-1.45, 1.19) | 0.0 (0.850) |
| Temporal | N=204 (317) | -0.11 (-1.60, 1.38) | 0.0 (0.888) | 0.07 (-1.20, 1.33) | 0.0 (0.920) |
| Superior | N=203 (316) | -0.15 (-1.81, 1.50) | 0.0 (0.857) | -0.16 (-1.76, 1.43) | 0.0 (0.839) |
| Nasal | N=204 (317) | -0.11 (-1.71, 1.48) | 0.0 (0.891) | -0.25 (-1.75, 1.26) | 0.0 (0.750) |
| Inferior | N=204 (316) | -0.31 (-2.14, 1.52) | 0.0 (0.739) | -0.28 (-1.89, 1.34) | 0.0 (0.739) |
| **Avanti Vessel Density (%)** |  |  |  |  |  |
| Whole image | N=204 (317) | -0.35 (-0.87, 0.17) | 0.7 (0.191) | -0.54 (-0.97, -0.10) | 2.3 (0.016) |
| Parafovea | N=204 (317) | -0.42 (-0.97, 0.13) | 0.9 (0.138) | -0.63 (-1.09, -0.18) | 2.8 (0.007) |
| Temporal | N=204 (317) | -0.38 (-0.95, 0.18) | 0.7 (0.183) | -0.49 (-0.96, -0.01) | 1.5 (0.046) |
| Superior | N=203 (316) | -0.31 (-0.88, 0.26) | 0.4 (0.293) | -0.59 (-1.09, -0.09) | 2.0 (0.022) |
| Nasal | N=204 (317) | -0.48 (-1.05, 0.08) | 1.1 (0.096) | -0.75 (-1.24, -0.25) | 3.2 (0.004) |
| Inferior | N=204 (316) | -0.57 (-1.26, 0.11) | 1.0 (0.102) | -0.78 (-1.36, -0.21) | 2.7 (0.008) |
| **Spectralis Choroid (μm)** |  |  |  |  |  |
| Global | N=247 (400) | -10.95 (-14.96, -6.94) | 8.9 (<0.001) | -15.17 (-18.96, -11.38) | 17.3 (<0.001) |
| Inner Ring | N=247 (400) | -11.51 (-16.18, -6.84) | 7.5 (<0.001) | -16.04 (-20.49, -11.58) | 14.6 (<0.001) |
| Outer Ring | N=247 (400) | -10.81 (-14.70, -6.92) | 9.2 (<0.001) | -14.97 (-18.64, -11.30) | 17.9 (<0.001) |
| Inner Temporal | N=247 (400) | -8.05 (-12.53, -3.57) | 4.1 (<0.001) | -12.33 (-16.64, -8.02) | 9.6 (<0.001) |
| Inner Superior | N=247 (400) | -12.56 (-17.72, -7.40) | 7.3 (<0.001) | -17.30 (-22.26, -12.34) | 13.8 (<0.001) |
| Inner Nasal | N=247 (400) | -16.87 (-22.38, -11.36) | 11.1 (<0.001) | -21.73 (-27.06, -16.41) | 17.9 (<0.001) |
| Inner Inferior | N=247 (400) | -9.28 (-14.19, -4.37) | 4.5 (<0.001) | -14.19 (-18.87, -9.51) | 10.8 (<0.001) |
| Outer Temporal | N=247 (400) | -6.56 (-10.30, -2.83) | 4.0 (<0.001) | -11.03 (-14.47, -7.60) | 11.8 (<0.001) |
| Outer Superior | N=247 (400) | -13.07 (-17.89, -8.24) | 8.9 (<0.001) | -17.45 (-22.12, -12.78) | 15.5 (<0.001) |
| Outer Nasal | N=247 (400) | -14.37 (-18.70, -10.04) | 12.8 (<0.001) | -18.10 (-22.34, -13.86) | 19.3 (<0.001) |
| Outer Inferior | N=247 (400) | -10.15 (-14.80, -5.50) | 6.0 (<0.001) | -14.92 (-19.32, -10.51) | 13.1 (<0.001) |

\*Linear mixed models slope estimates (with 95% confidence intervals) from univariable and multivariable models adjusted for age and VFMD. <sup>a</sup>R<sup>2</sup> reported as a percentage
Abbreviations: GCC; Ganglion cell complex, GCIPL; Ganglion cell inner plexiform layer, RNFL; Retinal nerve fiber layer, VFMD; Visual field mean deviation

### Associations with Severity of Glaucoma (Visual Field MD)

In multivariable models adjusted for age and axial length we found relatively strong associations between thinner global and sectoral GCIPL measures and worse VFMD (R^2^ range: 14.0%-38.1%, all p<0.001). Thinner mRNFL was also significantly associated with worse VFMD in all sectors (R^2^ range: 3.1%-23.8%, all p<0.001) with exception of the inner nasal and temporal sectors (p>0.285). Thinner global and sectoral Spectralis and Avanti GCC measures were significantly associated with worse VFMD (R^2^ range: from 20.2% to 37.0% and 18.6% to 35.4%, all p<0.001, respectively). In addition, we found a relatively strong association between lower macular vessel density and worse VFMD (R^2^ ranged from 20.3% to 33.2%, all p<0.001). Macular choroidal thickness was not associated with VFMD (Table 3).

**Table 3.**
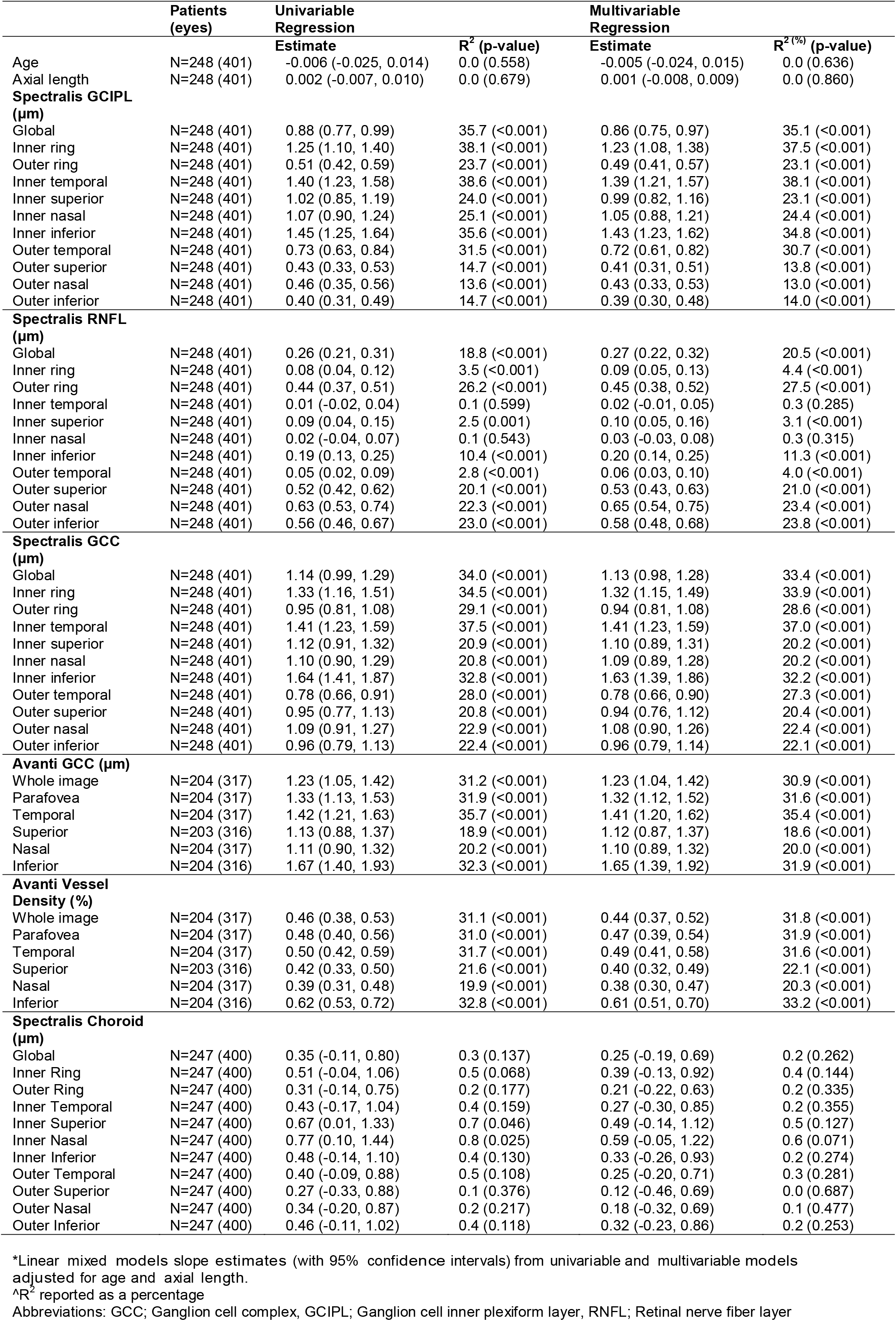
Ocular characteristics associations with Visual Field mean deviation

### Secondary Analysis of Differences by Axial Myopia Group

As a secondary analysis, we compared macular thickness- and vascular measurement differences across the three axial myopia groups and adjusted for age and VFMD. In general, mRNFL was thickest in high myopic eyes in all sectors, while mCT was significantly thinner in all sectors in high myopic eyes. The pattern of other macular thickness and vessel density measurements were less consistent across the three axial myopia groups (Supplemental Table 1 and Figures 2A-2F.)

Specifically, global and sectoral GCC (both Spectralis and Avanti) and GCIPL thickness values were similar across myopic groups, except for the inner and outer nasal rings, and inferior outer ring GCIPL (all p≤0.033) (see Figures 2A, 2C and 2D). Compared to no and mild myopia groups, thicker mRNFL was generally found in high myopes globally and in specific sectors (age and VF adjusted MD global: p=0.031, global inner: p=0.051, global outer p=0.042, inner superior ring p=0.001 and outer superior ring (p=0.017) (Figure 2B). Parafoveal vessel density tended to be slightly higher in non-myopes compared to mild and high myopes, but only reached statistical significance in the nasal sector (mean [95% CI]); nasal vessel density in high myopes (42.7% [40.7%, 44.6%]) and non-myopes (44.5% [43.3%, 45.7%]) (p=0.011) (Figure 2E). Global and sectoral mean MCT was significantly thinner in high myopes compared to mild and non-myopes (all p<0.001, See Figure 2F).

## Discussion

The results of this work have implications for diagnosing glaucoma in the challenging patients with high myopia. Specifically, our results suggest that macula measurements can be useful measurements to diagnose and monitor glaucoma in myopic eyes as the GCIPL and GCC thinned with increasing severity of glaucoma but are not associated with axial length. Except for choroidal thickness, all other macula thickness measures obtained in this study were associated with the severity of glaucoma. Because ganglion cell-related macular thickness measurements are strongly associated with VFMD but do not vary with axial length, GCIPL and GCC show promise for detecting glaucoma in myopic eyes.

Because the macula is devoid of morphometric variations such as tilt and peripapillary atrophy, it’s diagnostic role in detecting glaucoma in highly myopic eyes is gaining more attention recently. Specifically, there is evidence that myopia can lead to a high rate of false-positives in the measurement of the peripapillary RNFL (pRNFL).^23^ Several studies have examined the diagnostic ability of GCIPL, pRNFL and GCC and reported that GCIPL thickness has been reported as superior ^8 9 24-27^ or comparable to pRNFL thickness^28^ for diagnosing glaucoma in myopic eyes. Shoji et al. found that GCC parameters had high diagnostic accuracy to detect glaucoma in highly myopic eyes and that the diagnostic ability was higher than that of the pRNFL.^8^ In another study, these authors reported that GCC parameters were not significantly related to refractive errors and had good accuracy to detect glaucoma in non-myopes and in high myopes.^9^ Similarly, Kim et al. determined that in highly myopic eyes, the accuracy of glaucoma detection based on the macular GCC thickness was comparable to that based on the pRNFL thickness.^29^ These findings and those of other studies^8 9 27^ are consistent with our results that GCIPL and GCC thickness measured using both Spectralis and Avanti showed no association between the GCIPL thickness and axial length suggesting GCIPL thickness is less sensitive to changes due to axial elongation. Moreover, in our study GCIPL and GCC measurements showed the strongest association with VFMD, suggesting that both are useful for measuring ganglion cell loss associated with glaucoma in both non-highly myopic and highly myopic eyes. These results are generalizable across instruments as both Spectralis GCC and Avanti GCC showed similar results.

High myopia is characterized by marked structural changes in the retina and choroid and the corresponding vasculature.^30 31^ With the introduction of the non-invasive technique, OCTA images can provide a microvascular map from different retinal layers. To the best of our knowledge this is the first study comparing both macular tissue thickness and vascular measurements in axial non-myopic, mild and high myopic glaucomatous eyes. In the current study, the macular vessel density showed a weak association with axial length and a moderate association with VFMD. Previous studies reported conflicting results. This inconsistency can be explained in part by differences in study populations and image acquisition and analysis protocols. For instance, we employed a 3 × 3 mm imaging area whereas Yang et al. employed a larger 6 × 6 mm area.^31^ A large scan size can be more sensitive to image artefacts but also may identify microvasculature dropout in outer regions.^31^ We found only a weak association between superficial macula vessel density and axial length but a moderate association between vessel density and VFMD. Our results suggest that although myopic changes might affect vessel density in the macula, effects due to glaucoma are much stronger as indicated by the stronger association to the VFMD and therefore may also be useful for monitoring glaucoma in myopic eyes.^32^

In terms of choroidal thickness, as axial length increased, the choroid thinned in all sectors. Previously reported results have demonstrated choroidal thinning in highly myopic eyes.^33-35^ Ho et al.^33^ reported that subfoveal choroidal thickness decreased by 6.20 µm for each diopter of myopia and was thinnest in the nasal sectors in all groups, which is similar to the distributions of non-axial myopes, mild axial myopes and high axial myopes in our study. The choroid is a highly vascular layer, supplied by the posterior ciliary arteries and provides the retinal photoreceptors and the retinal pigment epithelium with oxygen and nourishment.^36^ Our results did not show an association between choroidal thickness and VFMD which suggests that choroidal thickness likely is not a useful metric for differentiating glaucomatous from healthy eyes or for monitoring glaucomatous progression.

The current study has several limitations. First, individuals with high myopia were younger. As retinal tissue is known to thin in older eyes^37^ we adjusted for age and VFMD in all analyses. In addition, we compared the 3 axial myopic groups after age-matching and found similar results (data not shown) with respect to the pattern of the retinal and vascular measurements in the three groups. Second, it has been suggested that axial length might affect retinal vessel density measurements and lead to incorrect scaling in OCTA imaging, which should be taken into account when interpreting our results.^38^ Moreover, as vessel density measurements vary across instruments,^39^ these sVD results are not necessarily generalizable to macula vessel density measurements from other OCTA instruments or to macula deep layer vessel density measurements. In addition, axial elongation often leads to retinal layer segmentation errors and measurement failures. However, we reviewed the OCT images meticulously for segmentation errors and excluded data with uncorrectable segmentation failures. Finally, the sample size of the high myopic group was relatively small compared to the other two groups and the mean axial length was only 26.5 mm. We can therefore not generalize our results to eyes with longer axial length.

In conclusion, GCIPL and GCC thickness can be useful measurements to diagnose and monitor glaucoma in myopic eyes as they thinned with increasing severity of glaucoma but did not vary with axial length. Macular sVD may also be useful for detecting glaucoma in myopic eyes, however we found a weak association between vessel density and axial length which needs to be explored further.

## Data Availability

The DIGS dataset is available upon request after appropriate data use agreements are initiated.

## Funding

### Grant support

JR: German Research Foundation research fellowship grant recipient (RE 4155/1-1) and German Ophthalmological Society Grant

MC: K99EY030942

SM: Tobacco-Related Disease Research Program T31IP1511

RNW: National Eye Institute R01EY029058, an Unrestricted grant from Research to Prevent Blindness (New York, NY)

LMZ: National Eye Institute R01EY011008, R01EY019869, R01EY027510, P30EY022589

