## Supplemental Table 1 for "Macula structural and vascular differences in glaucoma eyes with and without high axial myopia"

|  | **No Myopia** | **Mild Myopia** | **High Myopia** | **Overall** | **P-value** (age and VFMD adjusted) |
| --- | --- | --- | --- | --- | --- |
| **Spectralis GCIPL (µm)** | 87 (146) | 125 (208) | 36 (47) | 248 (401) |  |
| Global | 62.7 (60.8, 64.6) | 60.8 (59.2, 62.4) | 61.1 (58.0, 64.2) | 61.5 (60.3, 62.8) | 0.314 (0.350) |
| Inner ring | 72.5 (69.8, 75.1) | 70.6 (68.4, 72.8) | 70.3 (66.0, 74.6) | 71.2 (69.6, 72.9) | 0.490 (0.571) |
| Outer ring | 52.9 (51.5, 54.3) | 51.0 (49.9, 52.2) | 52.0 (49.8, 54.2) | 51.9 (51.0, 52.7) | 0.1031 (0.0911 ) |
| Inner temporal | 65.9 (62.9, 68.8) | 64.9 (62.5, 67.4) | 67.3 (62.5, 72.2) | 65.6 (63.7, 67.4) | 0.643 (0.579) |
| Inner superior | 76.6 (73.9, 79.4) | 74.6 (72.3, 76.9) | 73.7 (69.2, 78.1) | 75.2 (73.5, 76.9) | 0.403 (0.328) |
| Inner nasal | 77.1 (74.3, 79.9) | 74.3 (71.9, 76.6) | 71.5 (67.0, 76.0) | 75.0 (73.2, 76.8) | 0.0932 (0.033^2^) |
| Inner inferior | 70.2 (67.1, 73.3) | 68.6 (66.0, 71.2) | 68.1 (63.0, 73.3) | 69.1 (67.2, 71.0) | 0.683 (0.858) |
| Outer temporal | 51.9 (50.2, 53.6) | 50.3 (48.8, 51.7) | 51.8 (49.0, 54.6) | 51.1 (50.0, 52.1) | 0.256 (0.422) |
| Outer superior | 52.4 (50.9, 53.9) | 51.0 (49.7, 52.2) | 51.1 (48.7, 53.5) | 51.5 (50.6, 52.4) | 0.298 (0.169) |
| Outer nasal | 58.2 (56.6, 59.9) | 56.2 (54.8, 57.6) | 55.3 (52.7, 58.0) | 56.9 (55.8, 57.9) | 0.091 (0.011^1,2^) |
| Outer inferior | 49.0 (47.6, 50.4) | 46.8 (45.6, 47.9) | 49.8 (47.6, 52.0) | 48.0 (47.1, 48.9) | 0.0061,3 (0.0091,3 ) |
| **Spectralis RNFL (µm)** | 87 (146) | 125 (208) | 36 (47) | 248 (401) |  |
| Global | 24.7 (23.9, 25.5) | 24.6 (23.9, 25.3) | 25.7 (24.4, 27.0) | 24.8 (24.3, 25.3) | 0.269 (0.0312,3 ) |
| Inner ring | 20.7 (20.1, 21.2) | 20.7 (20.3, 21.2) | 21.5 (20.6, 22.4) | 20.8 (20.5, 21.2) | 0.276 (0.0512,3 ) |
| Outer ring | 28.7 (27.6, 29.9) | 28.4 (27.5, 29.4) | 29.9 (28.1, 31.8) | 28.7 (28.0, 29.4) | 0.319 (0.0422,3 ) |
| Inner temporal | 18.9 (18.5, 19.4) | 18.7 (18.4, 19.1) | 18.9 (18.2, 19.6) | 18.8 (18.6, 19.1) | 0.693 (0.473) |
| Inner superior | 22.0 (21.2, 22.7) | 21.9 (21.3, 22.6) | 24.1 (22.8, 25.3) | 22.2 (21.7, 22.7) | 0.0072,3 (0.0012,3 ) |
| Inner nasal | 20.0 (19.2, 20.7) | 20.3 (19.7, 20.9) | 21.1 (19.9, 22.3) | 20.3 (19.8, 20.7) | 0.293 (0.0842 ) |
| Inner inferior | 21.7 (20.9, 22.4) | 22.0 (21.4, 22.7) | 22.0 (20.7, 23.2) | 21.9 (21.4, 22.4) | 0.770 (0.279) |
| Outer temporal | 19.0 (18.6, 19.5) | 19.1 (18.7, 19.4) | 19.5 (18.8, 20.2) | 19.1 (18.8, 19.4) | 0.487 (0.0492 ) |
| Outer superior | 30.3 (28.8, 31.8) | 29.3 (28.1, 30.6) | 32.5 (30.0, 34.9) | 30.1 (29.1, 31.0) | 0.0633 (0.0172,3 ) |
| Outer nasal | 38.0 (36.3, 39.8) | 37.8 (36.3, 39.2) | 38.8 (36.0, 41.6) | 38.0 (36.9, 39.1) | 0.781 (0.314) |
| Outer inferior | 27.6 (26.0, 29.1) | 27.6 (26.3, 28.9) | 28.8 (26.2, 31.3) | 27.7 (26.8, 28.7) | 0.692 (0.1252 ) |
| **Spectralis GCC (µm)** | 87 (146) | 125 (208) | 36 (47) | 248 (401) |  |
| Global | 87.4 (84.8, 89.9) | 85.4 (83.3, 87.6) | 86.8 (82.7, 91.0) | 86.3 (84.7, 87.9) | 0.460 (0.757) |
| Inner ring | 93.1 (90.2, 96.1) | 91.3 (88.8, 93.8) | 91.8 (86.9, 96.6) | 92.0 (90.2, 93.9) | 0.631 (0.906) |
| Outer ring | 81.6 (79.3, 83.9) | 79.5 (77.5, 81.4) | 81.9 (78.2, 85.6) | 80.6 (79.1, 82.0) | 0.246 (0.391) |
| Inner temporal | 84.8 (81.8, 87.8) | 83.6 (81.1, 86.1) | 86.2 (81.3, 91.2) | 84.4 (82.5, 86.2) | 0.594 (0.486) |
| Inner superior | 98.6 (95.4, 101.8) | 96.5 (93.8, 99.2) | 97.7 (92.5, 102.9) | 97.4 (95.4, 99.4) | 0.578 (0.805) |
| Inner nasal | 97.1 (93.9, 100.3) | 94.6 (91.9, 97.2) | 92.7 (87.6, 97.7) | 95.3 (93.2, 97.3) | 0.284 (0.255) |
| Inner inferior | 91.9 (88.3, 95.5) | 90.7 (87.6, 93.7) | 90.1 (84.0, 96.1) | 91.1 (88.8, 93.3) | 0.833 (0.943) |
| Outer temporal | 71.0 (69.0, 72.9) | 69.3 (67.7, 70.9) | 71.4 (68.2, 74.5) | 70.2 (68.9, 71.4) | 0.285 (0.423) |
| Outer superior | 82.7 (80.0, 85.4) | 80.3 (78.0, 82.6) | 83.6 (79.1, 88.0) | 81.6 (79.9, 83.3) | 0.224 (0.381) |
| Outer nasal | 96.2 (93.2, 99.2) | 94.0 (91.5, 96.5) | 94.2 (89.4, 99.0) | 94.9 (93.0, 96.7) | 0.500 (0.697) |
| Outer inferior | 76.6 (73.9, 79.3) | 74.4 (72.2, 76.7) | 78.4 (74.0, 82.8) | 75.7 (74.0, 77.4) | 0.180 (0.158) |
| **Avanti GCC (µm)** | 69 (114) | 103 (164) | 32 (39) | 204 (317) |  |
| Whole image | 85.7 (82.7, 88.7) | 85.0 (82.5, 87.5) | 85.7 (81.0, 90.4) | 85.3 (83.5, 87.2) | 0.923 (0.993) |
| Parafovea | 91.1 (87.9, 94.3) | 90.0 (87.3, 92.7) | 91.0 (85.9, 96.0) | 90.5 (88.5, 92.5) | 0.842 (0.984) |
| Temporal | 82.8 (79.5, 86.0) | 82.5 (79.8, 85.1) | 84.6 (79.4, 89.8) | 82.8 (80.9, 84.8) | 0.757 (0.567) |
| Superior | 96.2 (92.6, 99.7) | 94.4 (91.4, 97.4) | 96.6 (91.0, 102.2) | 95.3 (93.1, 97.5) | 0.649 (0.816) |
| Nasal | 95.3 (91.9, 98.7) | 94.0 (91.2, 96.9) | 91.9 (86.7, 97.2) | 94.2 (92.1, 96.4) | 0.567 (0.268) |
| Inferior | 90.1 (86.2, 94.0) | 89.1 (85.8, 92.4) | 89.2 (82.8, 95.5) | 89.5 (87.0, 91.9) | 0.920 (0.991) |
| **Avanti Vessel Density (%)** | 69 (114) | 103 (164) | 32 (39) | 204 (317) |  |
| Whole image | 41.0 (39.9, 42.1) | 40.0 (39.1, 41.0) | 40.0 (38.2, 41.7) | 40.4 (39.7, 41.1) | 0.394 (0.147) |
| Parafovea | 44.0 (42.8, 45.2) | 42.8 (41.8, 43.8) | 42.8 (40.9, 44.7) | 43.2 (42.5, 44.0) | 0.270 (0.081) |
| Temporal | 41.8 (40.6, 43.0) | 41.1 (40.1, 42.1) | 41.3 (39.3, 43.2) | 41.4 (40.6, 42.1) | 0.676 (0.653) |
| Superior | 45.7 (44.5, 47.0) | 44.3 (43.3, 45.3) | 45.0 (43.0, 46.9) | 44.9 (44.1, 45.7) | 0.203 (0.0601 ) |
| Nasal | 44.5 (43.3, 45.7) | 42.9 (41.9, 43.9) | 42.7 (40.7, 44.6) | 43.5 (42.7, 44.2) | 0.0931 (0.0111,2 ) |
| Inferior | 43.9 (42.5, 45.4) | 42.7 (41.5, 44.0) | 42.0 (39.6, 44.3) | 43.1 (42.2, 44.0) | 0.288 (0.0822 ) |
| **Spectralis Choroid (µm)** | 87 (146) | 125 (208) | 35 (46) | 247 (400) |  |
| Global | 169.0 (160.9, 177.1) | 158.1 (151.1, 165.0) | 134.3 (122.6, 146.1) | 159.2 (153.3, 165.2) | <0.0011,2,3 (<0.001^1,2,3^) |
| Inner Ring | 183.5 (174.0, 192.9) | 172.8 (164.7, 180.8) | 145.1 (131.2, 159.0) | 173.3 (166.6, 180.1) | <0.0012,3 (<0.0011,2,3 ) |
| Outer Ring | 164.8 (156.9, 172.7) | 153.7 (147.0, 160.5) | 130.9 (119.5, 142.3) | 155.0 (149.3, 160.8) | <0.0011,2,3 (<0.001^1,2,3^) |
| Inner Temporal | 184.1 (175.0, 193.2) | 178.4 (170.7, 186.1) | 152.0 (137.8, 166.2) | 177.3 (171.2, 183.4) | <0.0012,3 (<0.001^2,3^) |
| Inner Superior | 186.0 (175.5, 196.6) | 169.2 (160.3, 178.1) | 143.6 (127.6, 159.6) | 172.3 (165.0, 179.6) | <0.0011,2,3 (<0.0011,2, ) |
| Inner Nasal | 176.0 (164.7, 187.2) | 157.0 (147.4, 166.5) | 123.1 (106.4, 139.7) | 159.9 (151.8, 167.9) | <0.0011,2,3 (<0.0011,2, ) |
| Inner Inferior | 188.6 (178.6, 198.5) | 187.2 (178.8, 195.6) | 156.9 (141.7, 172.0) | 184.0 (177.1, 190.8) | <0.0012,3 (<0.001^2,3^) |
| Outer Temporal | 173.2 (165.6, 180.8) | 166.1 (159.7, 172.5) | 147.5 (135.9, 159.2) | 166.4 (161.3, 171.6) | 0.0012,3 (<0.0011,2,3 ) |
| Outer Superior | 170.0 (160.1, 179.8) | 155.1 (146.7, 163.4) | 129.1 (114.3, 143.9) | 157.4 (150.5, 164.3) | <0.0011,2,3 (<0.001^1,2,3^) |
| Outer Nasal | 125.2 (116.3, 134.1) | 107.0 (99.5, 114.6) | 81.9 (68.7, 95.2) | 110.7 (104.3, 117.1) | <0.0011,2,3 (<0.001^1,2,3^) |
| Outer Inferior | 192.6 (183.2, 202.0) | 186.9 (178.9, 194.9) | 159.1 (145.0, 173.2) | 185.6 (179.0, 192.2) | <0.0012,3 (<0.001^2,3^) |

**Supplemental Table 1.** Macula parameters by axial myopia group

Results are presented as mean (95% confidence interval). Significance is determined by linear mixed models.

^1^No vs. Mild Myopia p< 0.05; ^2^No vs. High Myopia p< 0.05; ^3^Mild vs. High Myopia p< 0.05

Abbreviations: AL; Axial length, GCC; Ganglion cell complex, GCIPL; Ganglion cell inner plexiform layer, RNFL; Retinal nerve fiber layer, VFMD; Visual field mean deviation
